# Downregulation of Defensin genes in SARS-CoV-2 infection

**DOI:** 10.1101/2020.09.21.20195537

**Authors:** Mohammed M Idris, Sarena Banu, Archana B Siva, Ramakrishnan Nagaraj

**Affiliations:** CSIR-Centre for Cellular and Molecular Biology, Hyderabad 500007, India

**Keywords:** Defensin, COVID-19, gene regulation, qRT-PCR, SARS-CoV-2

## Abstract

Defensins, crucial components of the innate immune system, play a vital role against infection as part of frontline immunity. Association of SARS-CoV-2 infection with defensins has not been investigated till date. In this study, we have investigated the expression of defensin genes in the buccal cavity during COVID-19 infection. Nasopharyngeal/Oropharyngeal swab samples collected for screening SARS-CoV-2 infection were analyzed for the expression of major defensin genes by the quantitative real-time reverse transcription polymerase chain reaction, qRT-PCR. 40 SARS-CoV-2 infected positive and 40 negative swab samples were selected for the study. Based on the RT-PCR analysis involving gene specific primer for defensin genes, 10 defensin genes were found to be expressed in the Nasopharyngeal/Oropharyngeal cavity. Six defensin genes were further found to be significantly downregulated in SARS-CoV-2 infected patients as against the control, negative samples based on differential expression analysis. The genes significantly downregulated were defensin beta 4A, 4B, 106B, 107B, 103A and defensin alpha 1B. Downregulation of several defensin genes suggests that innate immunity provided by defensins is or may be compromised in SARS-CoV-2 infection resulting in progression of the disease caused by the virus. Upregulation of defensin gene expression and use of defensin peptides could be attractive therapeutic interventions.

## Introduction

The small cysteine rich cationic antibacterial proteins, defensins are crucial components of the innate immune system [1-6]. They are the first line of defense against bacterial, fungal and viral infections. Based on their pattern of disulfide bonding, mammalian defensins are classified into α, β, and θ subfamilies [1-6]. The α-defensins HNP1-4 are predominantly produced by neutrophils, stored in neutrophil azurophilic granules and released in large quantities upon neutrophil activation [3]. The neutrophil defensin genes DEFA1 and DEFA3 are subject to gene copy number variation, which could potentially relate to their level of expression and regulation of immune responses [7]. The β-defensins are expressed in a variety of epithelia especially in the airways [5]. Expression of the HBD1and HBD4 genes is essentially constitutive, whereas expression of HBD2 and 3 genes are inducible in response to various stimuli [5, 6]. Viruses, bacteria, microbial products, and pro-inflammatory cytokines, such as interleukin-1β (IL-1β) and tumor-necrosis factor (TNF), induce the expression of HBD2 and HBD3 in various cells [8-12]. In addition to its antimicrobial properties, DEFB4 (HBD2) is expressed in leukocytes and acts as a chemokine for cells of the adaptive immune response [13]. Defensins HD5 and HD6 are human enteric defensins and play important roles in gut immunity [15]. It is of interest to examine whether SARS-CoV-2 infection in humans is connected with defensin gene expression. SARS-CoV-2 has a heterogeneous pattern in exhibiting the symptoms associated with infection [16]. The varied pattern of symptoms might be associated with innate immune response of defensins. Effective defensin expression and clearance of the virus may result in mild or asymptomatic cases. A detailed expression analysis of human defensin genes could shed light on their role in modulating innate immune response, infection pattern and their association with the COVID-19 disease. In this study, we have examined the expression of defensin genes in cells from nasopharyngeal/oropharyngeal swab samples from normal subjects and patients tested positive for the COVID-19 disease by the real-time reverse transcription–polymerase chain reaction (qRT-PCR) method. We observed that several defensin genes were down regulated in patients.

## Methods

### Sample collection

Human nasopharyngeal/oropharyngeal swab samples in viral transport medium (VTM), received at CSIR-CCMB from Government Hospitals, Hyderabad, India for COVID-19 diagnosis were selected for the study after ethical approvals (Institutional Ethics Committee of CCMB - 82/2020). 40 SARS-CoV-2 positive and 40 SARS-CoV-2 negative samples were selected after real-time reverse transcription polymerase chain reaction (qRT-PCR) analysis for this study. The study also has approval of CCMB Institutional Biosafety Committee.

### SARS-CoV-2 diagnostics

Total RNA was extracted from the VTM swab samples using KingFisher™ Flex System (Thermofisher Scientific Inc., USA). The extracted RNA were analyzed for SARS-CoV-2 presence using single tube Reverse transcription-polymerase chain reaction (qRT-PCR) analysis using the Novel Coronavirus (2019-nCOV) RTPCR detection Kit (Shanghai Fosun Long March Medical Science CO. Ltd, China) for E gene and ORF gene of SARS-CoV-2 [17] according to manufacturer’s protocol. Quantstudio™ 5 (Thermofisher Scientific Inc., USA) was used to perform qRT-PCR. All the RT-PCR work carried out in a BSL-2 facility of CSIR-CCMB, Hyderabad, India.

### Defensin RT-PCR analysis

Gene specific primers were synthesized for a total of 18 various defensin genes and *GAPDH* housekeeping gene (Table 1). cDNA was synthesized from 50 μg of total RNA from each COVID positive and negative swab samples using iScript Advanced cDNA Synthesis Kit for RTPCR (BioRad, USA) following manufacturer’s protocol. qPCR analysis was performed for all the defensin genes and the housekeeping gene using TB Green Premix Ex Taq 11 (TliRNaseH Plus) kit (Takara, Japan) in Applied Biosystems ViiA(tm) 7 Real-Time PCR System (USA). The qPCR was performed using the following conditions - Initial denaturation at 95°C for 30 seconds; 40 cycle of 95°C (denaturation) for 15 seconds, 55 or 60°C (annealing) for 30 seconds and 72°C (extension) for 15 seconds; followed by melting curve analysis [18]. Amplification of gene specific single qPCR product was confirmed by performing gel electrophoresis of the amplified product in 2% agarose gel and melting curve analysis. The cycle threshold (Ct) value obtained from the analysis were used for differential expression analysis [18].

**Table 1:**
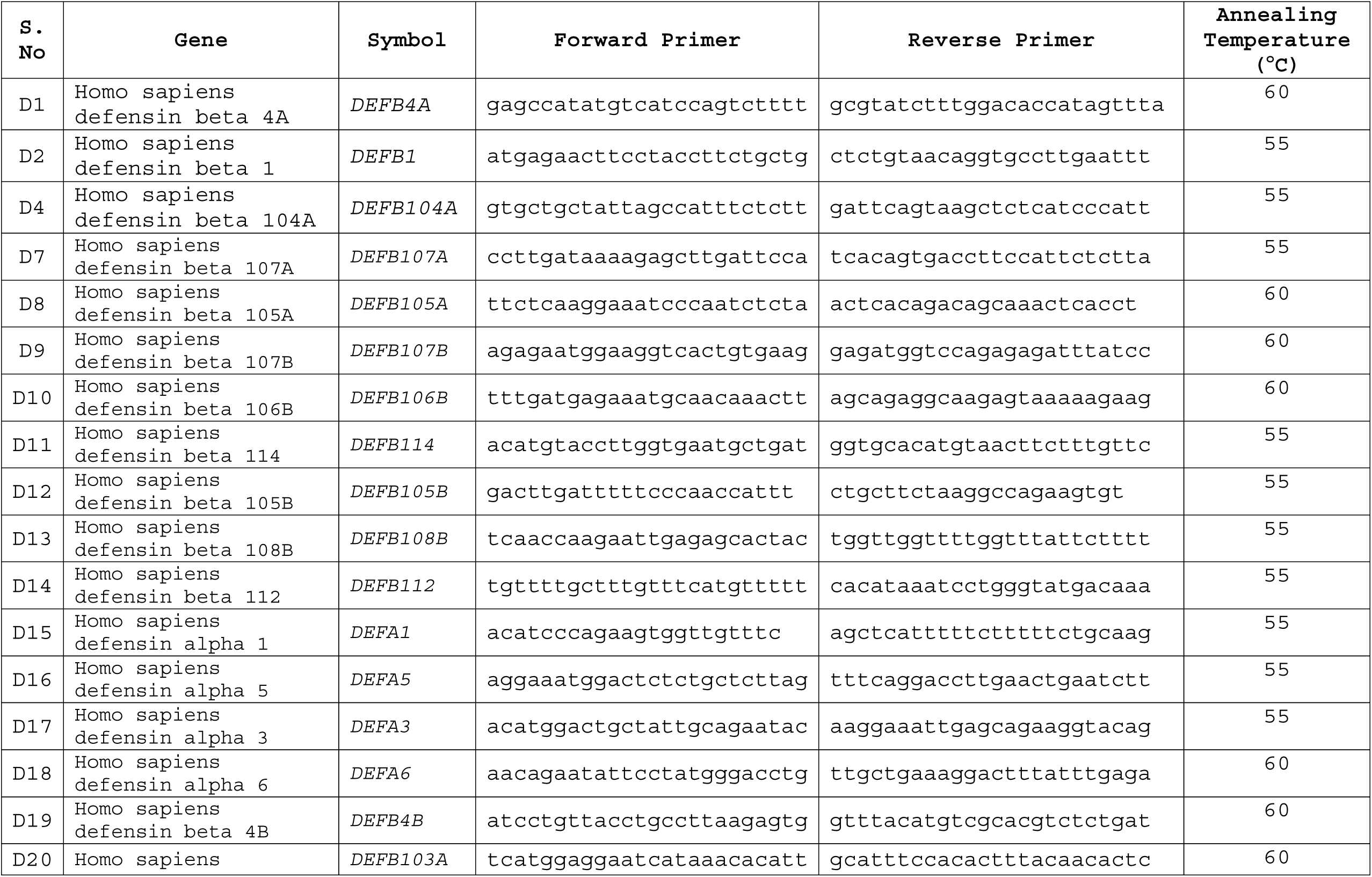
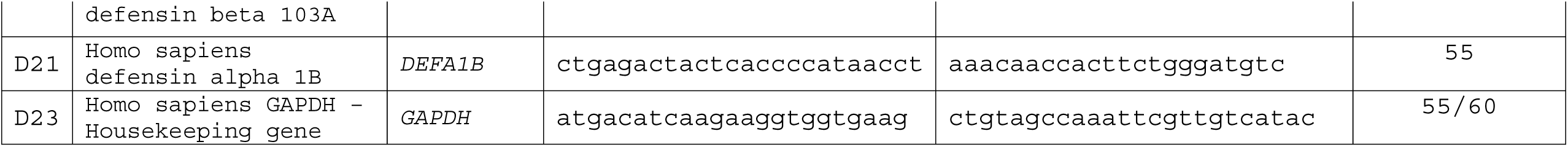
List of Defensin genes selected for the study and their forward and reverse Primer sequences with Annealing conditions.

### Differential expression analysis

The mean Ct value of each specific genes and housekeeping genes were calculated and their differential expression were analyzed based on the 2^delta Ct of target and housekeeping genes. The p values were calculated using standard t-test for all the observed Ct values. All the statistical analysis was performed using Microsoft Excel statistical software.

## Results and Discussion

### Samples

A total of 40 SARS-CoV-2 positive samples from the sample pool, having average Ct value of 23.2 for E gene and ORF gene of SARS-CoV-2 [17] were selected for the study. Similarly, 40 negative samples as controls for differential expression were selected from the same pool which tested negative to SARS-CoV-2 E and ORF gene.

### Expression of Defensin genes

The list of 18 defensin genes selected for the study are summarized in Table 1. They include both α- and β-defensin genes including isoforms. From these, 10 defensin genes showed expression at detectable limits in the selected Nasopharyngeal / Oropharyngeal swab samples, which were further selected for the differential expression analysis. The Ct values for the expression of defensin genes were selected in the range of 20.0 to 40.0 for all the genes including housekeeping genes. Undetectable range of amplification were excluded from the analysis. The mean Ct value of expression of defensin genes for positive and negative SARS-CoV-2 samples were analyzed separately (Table 2). Based on the expression analysis it was found that SARS-CoV-2 positive samples showed higher Ct values, in comparison to the SARS-CoV-2 negative samples for almost all the defensin genes, Differential expression analysis of the Ct value of each of the defensin genes against housekeeping *GAPDH* gene indicated that six defensin genes were significantly down regulated in the SARS-CoV-2 positive swab samples as against control SARS-CoV-2 negative swab samples.

**Table 2:** Expression level of Defensin genes for COVID-19 positive and negative samples.

| Gene Name | COVID Status | Ct Mean | Ct SEM | Differential Analysis | p Value | Significance |
| --- | --- | --- | --- | --- | --- | --- |
| COVID Status | Positive | 23.2 | 1.2 | - |  |  |
|  | Negative | - | - |  |  |  |
| Defensin beta 1 (*DEFB1*) | Positive | 32.5 | 0.4 | -0.14 | 0.09 | No |
|  | Negative | 32.1 | 0.2 |  |  |  |
| Defensin beta 4A (*DEFB4A*) | Positive | 29.9 | 0.4 | -0.72 | 0.04 | Yes |
|  | Negative | 28.9 | 0.3 |  |  |  |
| Defensin beta 105A (*DEFB105A*) | Positive | 31.2 | 0.2 | -0.13 | 0.19 | No |
|  | Negative | 30.9 | 0.3 |  |  |  |
| Defensin beta 107B (*DEFB107B*) | Positive | 29.8 | 0.4 | -1.00 | 0.02 | Yes |
|  | Negative | 28.5 | 0.3 |  |  |  |
| Defensin beta 106B (*DEFB106B*) | Positive | 29.0 | 0.4 | -1.33 | 0.01 | Yes |
|  | Negative | 27.5 | 0.3 |  |  |  |
| Defensin beta 4B (*DEFB4B*) | Positive | 29.6 | 0.4 | -0.95 | 0.02 | Yes |
|  | Negative | 28.4 | 0.3 |  |  |  |
| Defensin beta 103A (*DEFB103A*) | Positive | 29.1 | 0.4 | -1.12 | 0.01 | Yes |
|  | Negative | 27.8 | 0.3 |  |  |  |
| Defensin alpha 3 (*DEFA3*) | Positive | 29.7 | 0.5 | -0.67 | 0.11 | No |
|  | Negative | 28.7 | 0.4 |  |  |  |
| Defensin alpha 6 (*DEFA6*) | Positive | 29.9 | 0.5 | -0.48 | 0.07 | No |
|  | Negative | 29.2 | 0.4 |  |  |  |
| Defensin alpha 1B (*DEFA1B*) | Positive | 32.8 | 0.3 | -0.37 | 0.03 | Yes |
|  | Negative | 32.2 | 0.3 |  |  |  |
| *GAPDH* | Positive | 24.6 | 0.4 | - |  |  |
|  | Negative | 24.3 | 0.3 |  |  |  |

The β-defensin genes *DEFB4A, 107B, 106B, 4B* and *103A* were found significantly down regulated based on differential analysis against control for SARS-CoV-2 infection (Figure 1, Table 2). Interestingly, one α-defensin gene *DEFA1B* was also down regulated (Figure 1, Table 2). Genes corresponding to HD-5 and HD-6 were not affected as would be expected in nasopharyngeal/oropharyngeal samples. Of particular interest was the downregulation of both isoforms of the HBD-2 genes DEFB4A and DEFB4B. HBD-2 represents the human defensin that is produced by epithelial cells following contact with bacteria, viruses, or cytokines such as IL-1 and TNF-α [19]. HBD-2 appears to activate the primary antiviral innate immune response [20]. Although the nature of the regulation is not completely characterized, both the MAPK signal transduction pathway and the NF-κB transcription factor have been suggested to be involved. Also, the less investigated DEFB106 and DEFB107 were also downregulated.

**Figure 1:**
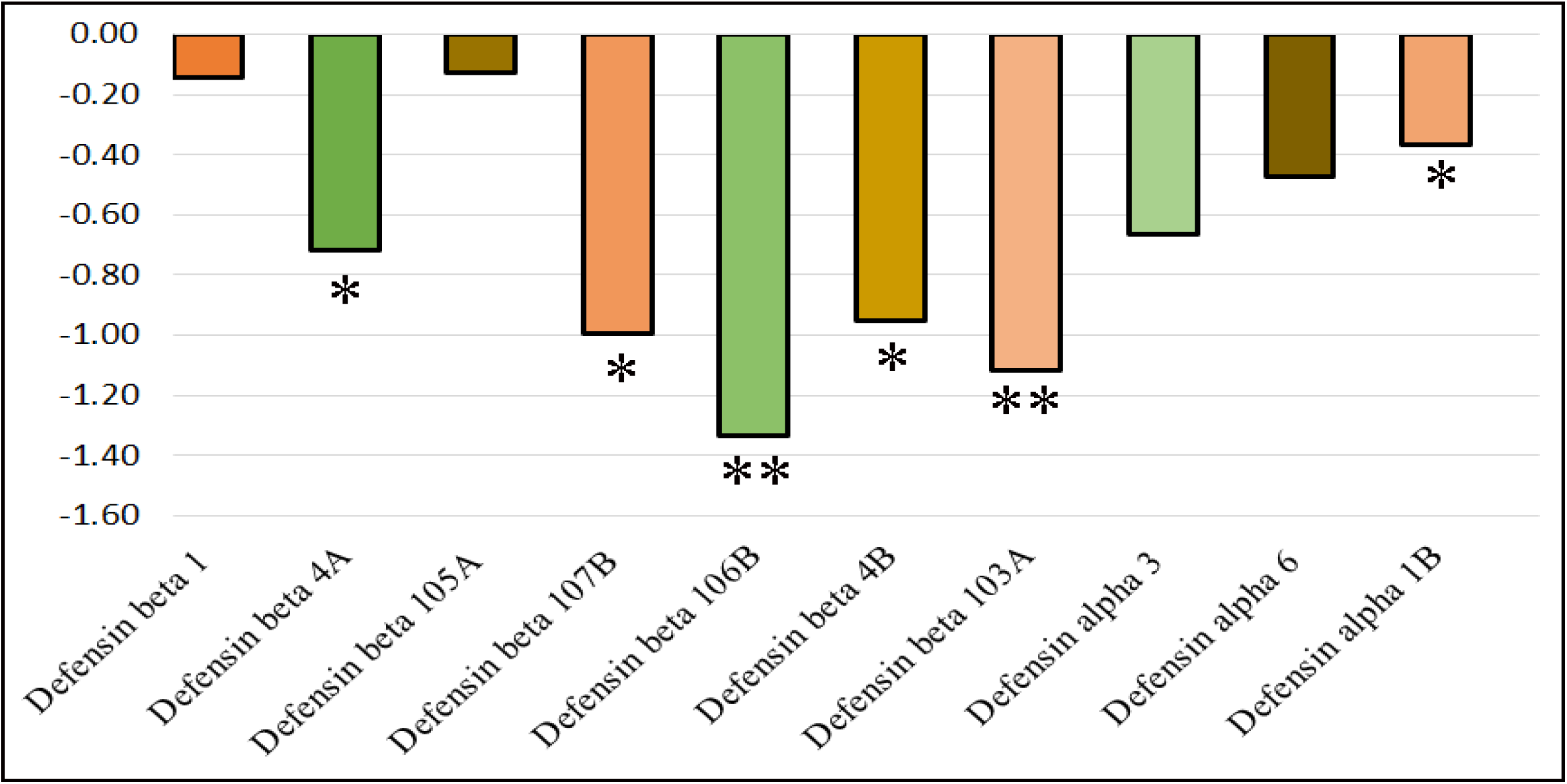
Differential expression level of Defensin genes in COVID-19 positive samples against COVID-19 negative samples. (* p = 0.05 to 0.01; ** p = <0.01).

The antiviral activities of human defensins has been the subject of extensive investigations [20-24]. Antiviral activity of defensins against HIV and other viruses suggest that defensins have a role in host-defense not only against bacteria. Their expression in several cell types and tissues [22] suggest that they can act as the first line of defense against viruses targeting various cell types. HD-5 has been shown to inhibit SARS-CoV-2 *in vitro* by binding to Angiotensin-converting enzyme-2 (ACE2), the cellular receptor for the virus [24]. Hence, the interest in exploring defensins for anti-viral therapeutics [21, 23].

The downregulation of several defensin genes suggests that innate immunity provided by defensins is compromised in SARS-CoV-2 infection resulting in progression of the disease caused by the virus. Also, virus-mediated downregulation of defensin expression could result in augmented colonization of the uppermost airway by bacteria resulting in lung infection. Association of defensin genes with SARS-CoV-2 infection suggests that upregulating defensin gene expression could be an attractive therapeutic intervention. Defensin peptides that are part of the full-length human defensins HBD-1-3 that exhibit antimicrobial activities could also conceivably act as effective antiviral agents for therapies against SARS-CoV-2 [25-27].

## Data Availability

All the datas mentioned in the manuscript are available.

## Acknowledgements

R.N. acknowledges the National Academy of Sciences, India for NASI Platinum Jubilee Senior Scientist Fellowship. R.N. is the recipient of JC Bose Fellowship from the Department of Science and Technology, India. Authors acknowledge funding from Council of Scientific and Industrial Research (CSIR), India.

## Notes

### Competing Interest Statement

The authors have declared no competing interest.

### Author Declarations

Institutional Ethics Committee of CCMB - 82/2020

## References

1. Oppenheim, J. J., Biragyn, A., Kwak, L. W., & Yang, D. Roles of antimicrobial peptides such as defensins in innate and adaptive immunity. Ann. Rheum. Dis. 2003, 62, ii17–ii21.

2. Ganz, T. The role of antimicrobial peptides in innate immunity. Integr. Comp. Biol. 2003, 43, 300–304.

3. Selsted, M. E., & Ouellette, A. J. Mammalian defensins in theantimicrobial immune response. Nat. Immunol. 2005, 6, 551−557.

4. Schneider, J. J., Unholzer, A., Schaller, M., Schafer-Korting, M., & Korting, H. C. Human Defensins. J. Mol. Med. 2005, 83, 587−595.

5. Pazgier, M., Hoover, D. M., Yang, D., Lu, W., & Lubkowski, J. Human beta-defensins. Cell. Mol. Life Sci. 2006, 63, 1294−1313.

6. Xu, D., & Lu, W. Defensins: A Double-Edged Sword in Host Immunity. Front Immunol. 2020, 11: 764

7. Mayumi, K., Holly, M. K., Diaz, K., & Smith. G.K. Defensins in Viral Infection and Pathogenesis. Annu Rev Virol. 2017, 4, 269–271.

8. Ding, J., Chou, Y.Y., & Chang, T.L. Defensins in viral infections. J. Innate Immun. 2009, 1, 413–420.

9. Klotman, M. E., & Chang, T. L. Defensins in innate antiviral immunity. Nat. Rev. Immunol. 2006, 6, 447–456.

10. Wilson, S. S., Wiens, M. E., Holly, M. K., & Smith, J. G. Defensins at the Mucosal Surface: Latest Insights into Defensin-Virus Interactions. J. Virol. 2016, 90, 5216–5218.

11. Reddick, L. E., & Alto, N. M. Bacteria fighting back: how pathogens target and subvert the host innate immune system. Mol.Cell 2014, 54, 321−328.

12. Semple, F., & Dorin J. R. ß-Defensins: multifunctional modulators of infection, inflammation and more? J Innate Immun. 2012,4(4):337–348.

13. Yang, D., Chertov O, Bykovskaia S. N., Chen, Q., Buffo, M. J., Shogan, J., Anderson, M., Schröder, J. M., Wang, J. M., Howard, O. M., & Oppenheim, J. J. Beta-defensins: linking innate and adaptive immunity through dendritic and T cell CCR6.

14. Findlay, F., Proudfoot, L., Stevens, C., & Barlow, P.G. Cationic host defense peptides, novel antimicrobial therapeutics against Category A pathogens and emerging infections. Pathog. Glob. Health 2016, 110, 137–147.

15. Clevers, H. C., & Bevins, C. L. Paneth cells: maestros of the small intestinal crypts. Annu Rev Physiol. (2013) 75:289–311

16. Cao, X. COVID-19: immunopathology and its implications for therapy. Nature Rev Immunol (2020) 20, 269–270.

17. Chu, D. K. W., Pan, Y., Cheng, S. M. S., Hui, K. P. Y., Krishnan, P., Liu, Y., Ng, D. Y. M., Wan, C. K. C., Yang, P., Wang, Q., Peiris, M., & Poon, L. L. M. (2020). Molecular diagnosis of a novel coronavirus (2019-nCoV) causing an outbreak of pneumonia. Clin Chem. 66, 549–555

18. Saxena, S., Singh, S. K., Meena Lakshmi, M. G., Meghah, V., Bhatti, B., Brahmendra Swamy, C. V., Sundaram, C. S., & Idris, M. M. Proteomics analysis of zebrafish caudal fin regeneration. Molecular and Cellular Proteomics 2012, 11(6):M111.014118.

19. O’Neil, D. A. Regulation of expression of β-defensins: endogenous enteric peptide antibiotics. Molecular Immunology 40 (2003) 445–450

20. Kim, J., Yang, Y.L. Jang, S-E, & Yong-Suk Jang, Y-S. Human β-defensin 2 plays a regulatory role in innate antiviral immunity and is capable of potentiating the induction of antigen specific immunity. Virology Journal (2018) 15:124

21. Park, M.S., Kim, J.I., Lee, I., Park, S., Bae, J.Y., & Park, M.S. Towards the Application of Human Defensins as Antivirals. Biomol. Ther. 2018, 26, 242–254.

22. Pace, B. T., Lackner, A. A., Porter, E., & Pahar, B. The Role of Defensins in HIV Pathogenesis. Mediat. Inflamm.2017, 2017, 5186904.

23. Ahmed, A., Siman-Tov, G., Hall, G., Bhalla, N., & Narayanan, A. Human Antimicrobial Peptides as Therapeutics for Viral Infections. Viruses 2019, 11, 704–730.

24. Wang, C., Wang, S., Li, D., Wei, D-Q Zhao. J., & Wang, J. Human Intestinal Defensin 5 Inhibits SARS-CoV-2 Invasion by Cloaking ACE2. Gastroenterology 2020. In press.

25. Hoover, D. M., Wu, Z., Tucker, K., Lu, W., & Lubkowski, J. Antimicrobial characterization of human beta-defensin 3 derivatives. Antimicrob. Agents Chemother. 2003, 47, 2804−2809.

26. Krishnakumari, V., Singh, S., & Nagaraj, R. Antibacterial activities of synthetic peptides corresponding to the carboxy-terminal region of human beta-defensins 1-3. Peptides 2006, 27, 2607−2613.

27. Krishnakumari, V., Guru, A., Adicherla, H., & Nagaraj, R. Effects of increasing hydrophobicity by N-terminal myristoylation on the antibacterial and hemolytic activities of the C-terminal cationic segments of human-beta-defensins 1-3. Chem. Biol. Drug Des. 2018, 92, 1504−1513.

